# Radiogenomic profiling of prostate tumors prior to external beam radiotherapy converges on a transcriptomic signature of TGF-β activity driving tumor recurrence

**DOI:** 10.1101/2023.05.01.23288883

**Authors:** Anson T. Ku, Uma Shankavaram, Shana Y. Trostel, Hong Zhang, Houssein A. Sater, Stephanie A. Harmon, Nicole V. Carrabba, Yang Liu, Bradford J. Wood, Peter A. Pinto, Peter L. Choyke, Radka Stoyanova, Elai Davicioni, Alan Pollack, Baris Turkbey, Adam G. Sowalsky, Deborah E. Citrin

## Abstract

**Background:** Patients with localized prostate cancer have historically been assigned to clinical risk groups based on local disease extent, serum prostate specific antigen (PSA), and tumor grade. Clinical risk grouping is used to determine the intensity of treatment with external beam radiotherapy (EBRT) and androgen deprivation therapy (ADT), yet a substantial proportion of patients with intermediate and high risk localized prostate cancer will develop biochemical recurrence (BCR) and require salvage therapy. Prospective identification of patients destined to experience BCR would allow treatment intensification or selection of alternative therapeutic strategies.

**Methods:** Twenty-nine individuals with intermediate or high risk prostate cancer were prospectively recruited to a clinical trial designed to profile the molecular and imaging features of prostate cancer in patients undergoing EBRT and ADT. Whole transcriptome cDNA microarray and whole exome sequencing were performed on pretreatment targeted biopsy of prostate tumors (n=60). All patients underwent pretreatment and 6-month post EBRT multiparametric MRI (mpMRI), and were followed with serial PSA to assess presence or absence of BCR. Genes differentially expressed in the tumor of patients with and without BCR were investigated using pathways analysis tools and were similarly explored in alternative datasets. Differential gene expression and predicted pathway activation were evaluated in relation to tumor response on mpMRI and tumor genomic profile. A novel TGF-β gene signature was developed in the discovery dataset and applied to a validation dataset.

**Findings:** Baseline MRI lesion volume and *PTEN*/*TP53* status in prostate tumor biopsies correlated with the activation state of TGF-β signaling measured using pathway analysis. All three measures correlated with the risk of BCR after definitive RT. A prostate cancer-specific TGF-β signature discriminated between patients that experienced BCR vs. those that did not. The signature retained prognostic utility in an independent cohort.

**Interpretation:** TGF-β activity is a dominant feature of intermediate-to-unfavorable risk prostate tumors prone to biochemical failure after EBRT with ADT. TGF-β activity may serve as a prognostic biomarker independent of existing risk factors and clinical decision-making criteria.

**Funding:** This research was supported by the Prostate Cancer Foundation, the Department of Defense Congressionally Directed Medical Research Program, National Cancer Institute, and the Intramural Research Program of the NIH, National Cancer Institute, Center for Cancer Research.

## INTRODUCTION

Localized prostate cancer is a heterogenous clinical entity for which numerous clinical and pathologic prognostic factors have been identified. Combining clinicopathologic variables, such as pretreatment PSA, Gleason Score, local tumor stage, and percent of biopsy cores with tumor has been used to define several distinct prognostic risk groups that inform treatment recommendations. Despite this useful framework, it remains challenging to predict which patients subjected to similar potentially curative treatments within or across risk groups are destined to experience local cancer recurrence or develop metastatic, incurable prostate cancer. Prospective identification of those patients at greatest risk of recurrence with initial treatment for localized prostate cancer would provide an opportunity for intensified treatment or alternative therapy.

Treatment with radiotherapy alone or in combination with androgen deprivation therapy (ADT) is highly effective as a curative treatment option for patients with localized prostate cancer, yet in a recent meta-analysis with 11 years median follow-up, local and distant recurrences were estimated to occur in 13% and 21% of high-risk patients, and 7.2% and 7.2% of intermediate-risk patients, respectively [1]. Efforts to improve prognostication in prostate cancer patients receiving radiotherapy with or without ADT have centered around prognostic genomic classifiers that were developed to predict for risk of metastases and recurrence in cohorts of patients treated with prostatectomy. Although these signatures have also been demonstrated to yield strong prognostic utility in patients receiving radiotherapy, it is plausible that the biological events responsible for recurrence in the setting of radiotherapy with or without ADT may vary from those that drive recurrence after prostatectomy, and as a result, that the optimal genomic classifier to predict for recurrence may vary between these distinct therapies.

Multiparametric magnetic resonance imaging (mpMRI) consisting of T2-weighted (T2W), diffusion-weighted (DWI), and dynamic contrast-enhanced (DCE) imaging provides superior soft tissue contrast within the pelvis compared to other available anatomic imaging modalities, while simultaneously providing functional assessment of visualized prostate lesions. Thus, mpMRI has played an increasing role in the diagnosis of prostate cancer through the use of MRI-US fusion guided biopsies, and in treatment of prostate cancer through use of focal tumor treatment approaches and focal dose escalation strategies for radiation.

Given the individual prognostic benefits of genomic classifiers and mpMRI to identify clinically significant prostate cancer, we conducted a prospective natural history study to profile the molecular and imaging features of prostate tumors undergoing EBRT plus ADT. Here, we present an integrated radiologic, genomic and transcriptomic analysis of 60 biopsies obtained from 43 MRI-visible prostate tumors in 29 patients. Pretreatment biopsies were subjected to whole-exome sequencing and whole-transcriptome cDNA microarrays. We observed that baseline MRI lesion volume and *PTEN*/*TP53* status correlate with the activation state of TGF-β signaling measured using pathway analysis, and all three measures correlated with the risk of biochemical recurrence after definitive RT. A novel prostate cancer-specific TGF-β signature discriminated between patients destined to experience biochemical failure vs. those that did not, and the signature retained this prognostic capacity in an independent cohort. Thus, our results indicate that TGF-β activity is a dominant features of intermediate-to-unfavorable risk prostate tumors prone to biochemical failure.

## RESULTS

### Baseline and posttreatment tumor volumes are prognostic of biochemical recurrence

The overall goal of this study was to identity radiologic and genomic features that are associated with biochemical recurrence following definitive EBRT plus ADT for intermediate and high risk prostate cancer. Of all patients enrolled in this prospective natural history study, we selected 29 study subjects with the longest follow-up duration (median follow-up of 91 months at the time of database lock) for whom sufficient biopsy tissue was available for molecular analyses (**Table 1**). Up to 4 lesions per patient were individually measured by mpMRI for a total of 43 lesions (excluding seminal vesicles, which were separately contoured where possible), both prior to initiation of therapy (all 29 patients) and six months after RT (28 patients). At the time of database lock, 5 patients had experienced biochemical recurrence. When stratified by clinical outcome, baseline mpMRI volume across all visible lesions was significantly greater in patients who experienced BCR (**Fig. 1A)**. Six months after RT, almost every lesion demonstrated a reduction in volume (**Fig. 1B)**. Similar to baseline, posttreatment mpMRI volume was significantly greater in patients who experienced BCR (**Fig. 1C**). However, the percentage change per lesion was not statistically significant between lesions from patients with BCR compared to lesions in patients without BCR, indicating that response at the lesion level was not predictive (**Fig. 1D**).

**Table 1.**
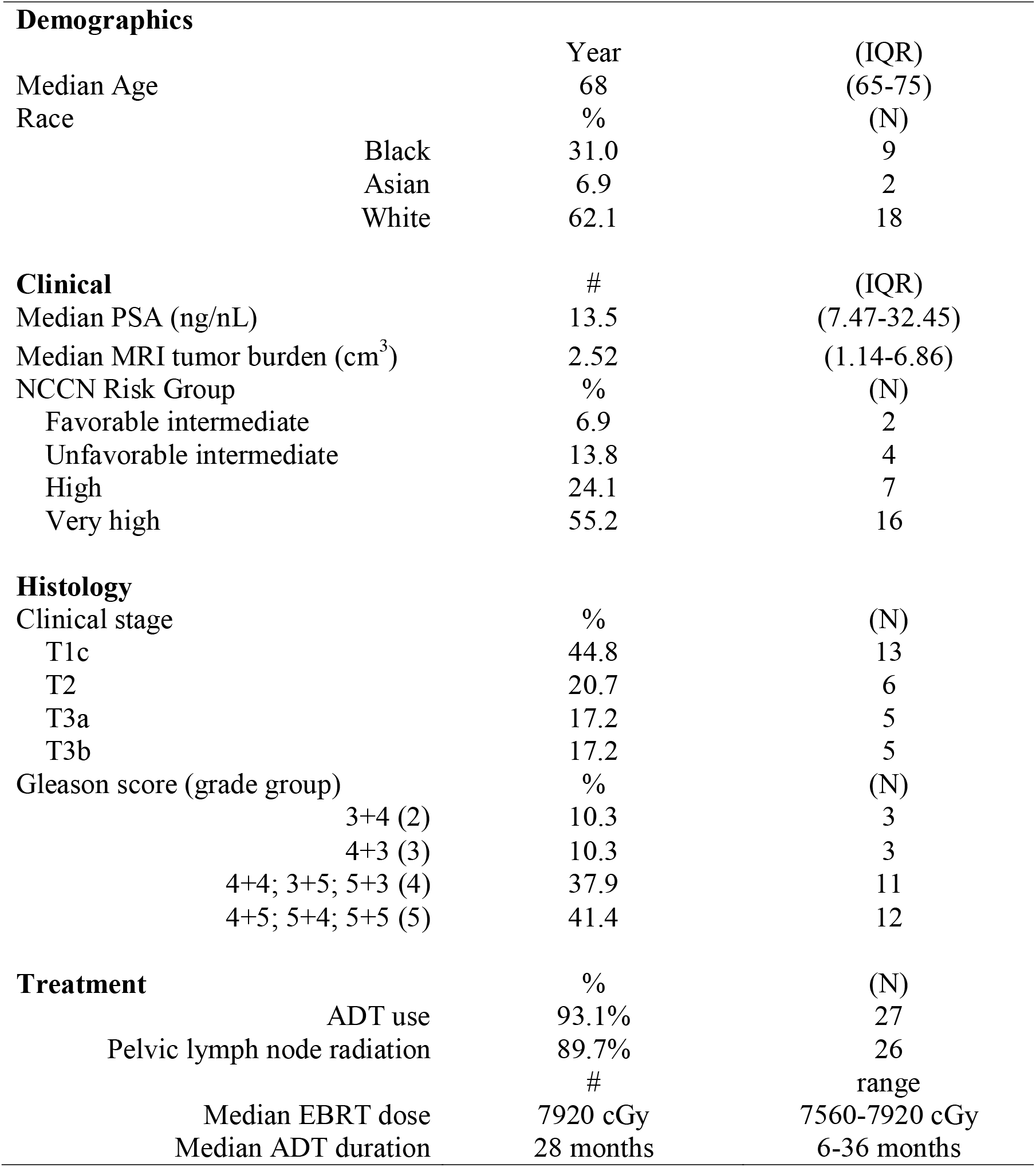
Patient Characteristics

| **Demographics** |  |  |
| --- | --- | --- |
|  | Year | (IQR) |
| Median Age | 68 | (65-75) |
| Race | % | (N) |
| Black | 31.0 | 9 |
| Asian | 6.9 | 2 |
| White | 62.1 | 18 |
| **Clinical** |  |  |
|  | # | (IQR) |
| Median PSA (ng/nL) | 13.5 | (7.47-32.45) |
| Median MRI tumor burden (cm<sup>3</sup>) | 2.52 | (1.14-6.86) |
| NCCN Risk Group | % | (N) |
| Favorable intermediate | 6.9 | 2 |
| Unfavorable intermediate | 13.8 | 4 |
| High | 24.1 | 7 |
| Very high | 55.2 | 16 |
| **Histology** |  |  |
| Clinical stage | % | (N) |
| T1c | 44.8 | 13 |
| T2 | 20.7 | 6 |
| T3a | 17.2 | 5 |
| T3b | 17.2 | 5 |
| Gleason score (grade group) | % | (N) |
| 3+4 (2) | 10.3 | 3 |
| 4+3 (3) | 10.3 | 3 |
| 4+4; 3+5; 5+3 (4) | 37.9 | 11 |
| 4+5; 5+4; 5+5 (5) | 41.4 | 12 |
| **Treatment** |  |  |
|  | % | (N) |
| ADT use | 93.1% | 27 |
| Pelvic lymph node radiation | 89.7% | 26 |
|  | # | range |
| Median EBRT dose | 7920 cGy | 7560-7920 cGy |
| Median ADT duration | 28 months | 6-36 months |

**Figure 1.**
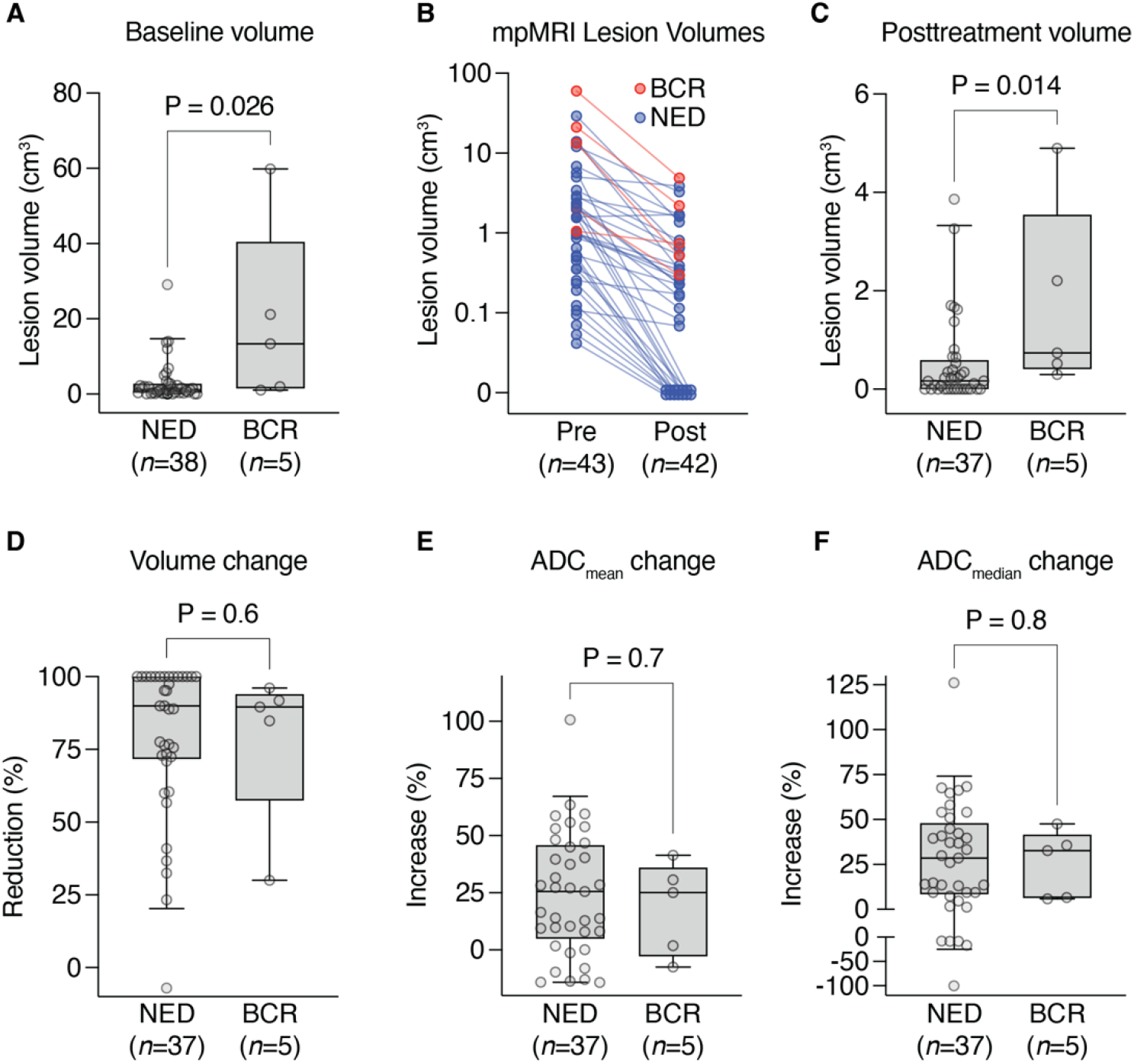
mpMRI features of radiation response. **(A)** Baseline lesion volumes of MRI-visible lesions for NED versus BCR patients. P = 0.026 by Mann-Whitney *U* test. **(B)** Change in MRI lesion volume from baseline to repeat MRI six months after RT. Lesions that had resolved completely were imputed to volume of 0 cm^3^. **(C)** Posttreatment lesion volumes of MRI-visible lesions for NED versus BCR patients. P = 0.014 by Mann-Whitney *U* test. **(D)** Per-lesion volume change, plotted as percent reduction, for NED versus BCR patients. P = 0.6 by Mann-Whitney *U* test. **(E)** Per-lesion change in mean ADC, plotted as percent increase, for NED versus BCR patients. P = 0.6 by Mann-Whitney *U* test. **(F)** Per-lesion change in median ADC, plotted as percent increase, for NED versus BCR patients. P = 0.8 by Mann-Whitney *U* test. For (**A** and **C-F**), boxes depict 25^th^ to 75^th^ percentile with line at median; whiskers extend to 5^th^ and 95^th^ percentile.

Previously, lower pretreatment ADC values in the index lesion has been demonstrated to predict for biochemical failure after radiotherapy [2, 3]. However, we did not observe this in our study, as neither the pretreatment ADC mean or median, post treatment ADC mean or median, nor percent change in ADC mean or median values (**Figs. 1E** and **1F**, respectively) were associated with outcome.

### Association of transcriptomic analyses with clinical outcomes

We next performed an integrative genomic analysis using biopsies targeted to and acquired from mpMRI-visible lesions using MRI/ultrasound-fusion guidance. Up to three biopsies were acquired per lesion for a total of 60 biopsies. RNA and DNA were extracted and used to perform microarray-based gene expression profiling (Affymetrix Human Exon array) and whole-exome sequencing, respectively (**Fig. 2**).

**Figure 2.**
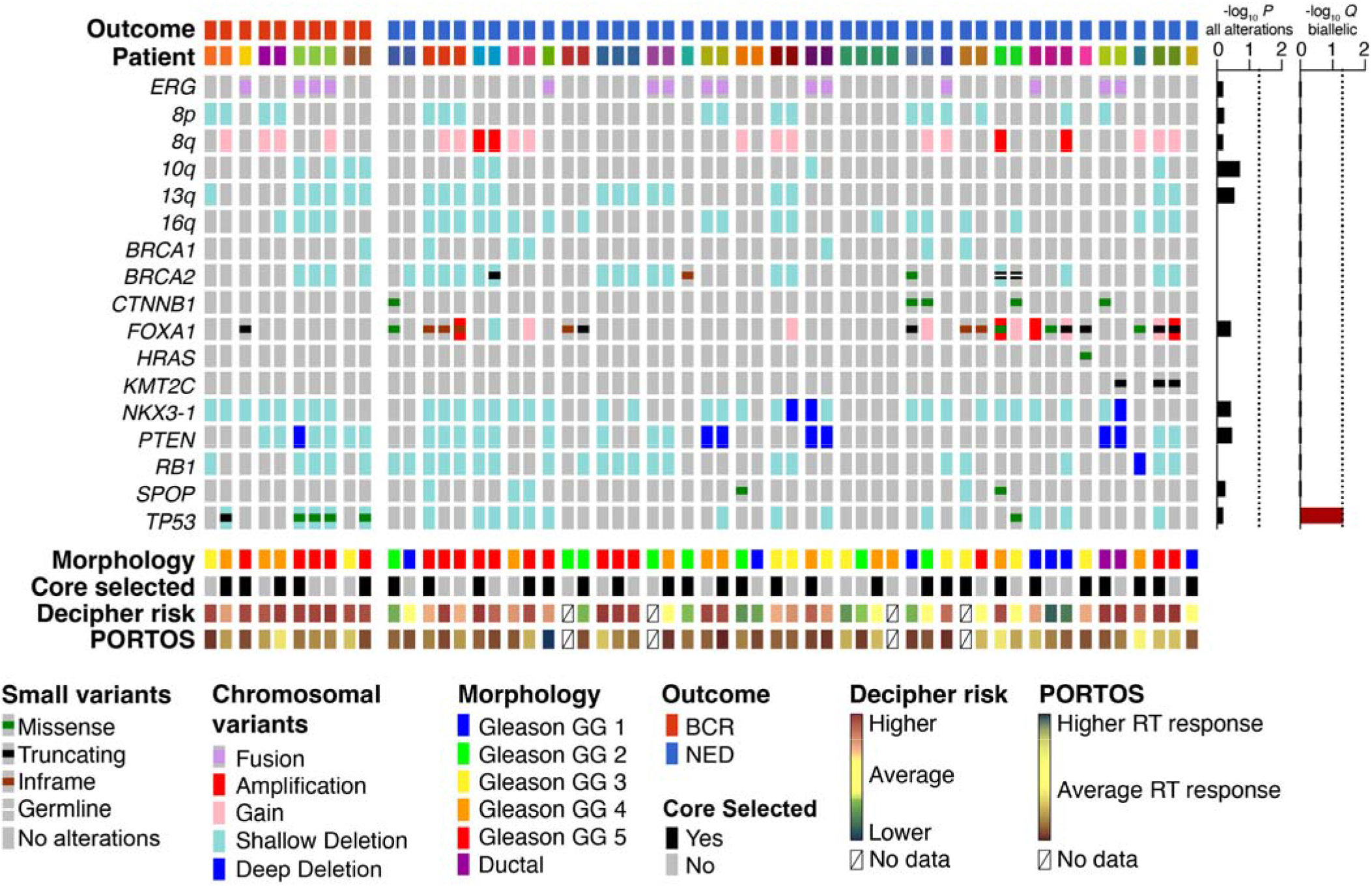
Molecular landscape of baseline prostate tumors prior to undergoing RT. Up to four biopsies were acquired per patient and subjected to whole-exome sequencing and transcriptome profiling on the Affymetrix Human Exon 1.0 array. Samples are grouped by outcome and separated by patient. Alterations in previously-identified prostate cancer MutSig genes are shown. From each case, a single biopsy was selected using predefined criteria, consisting of the most adverse phenotypes, for patient-level prognostication. Alteration enrichments were determined using a two-sided Fisher’s exact test with (Q) or without (P) Benjamini-Hochberg correction.

The microarray approach was chosen specifically due to its widespread use as a validated genomic classifier for predicting prostate cancer metastasis after prostatectomy, although its utility to predict recurrence after RT is unknown. Affymetrix CEL files were processed through the Decipher Genomic Resource Information Database to obtain classifier scores. Using pre-defined criteria for applying the Decipher classifier to cases with more than one biopsy (see Methods), we selected a single representative biopsy for per-patient analyses. Using validated cut-points for risk, there was no significant enrichment for higher risk classification in BCR vs. NED patients (P = 0.13, Cochran-Armitage test for trend). However, Decipher scores (measured continuously) were positively associated with both baseline (**Fig. 3A)** and posttreatment (**Fig. 3B**) patient-level tumor volumes, consistent with our observation of a positive correlation between baseline and post-treatment MRI tumor volumes (**Supplementary Fig. 1)**. Only the correlation with patient-level baseline tumor volumes was statistically significant. Surprisingly, application of the Post-Operative Radiation Therapy Outcomes Score (PORTOS) predicted only a single case to exhibit a superior response to RT; no significant association between PORTOS (measured continuously) and tumor volumes were observed (**Supplementary Fig. 2**).

**Figure 3.**
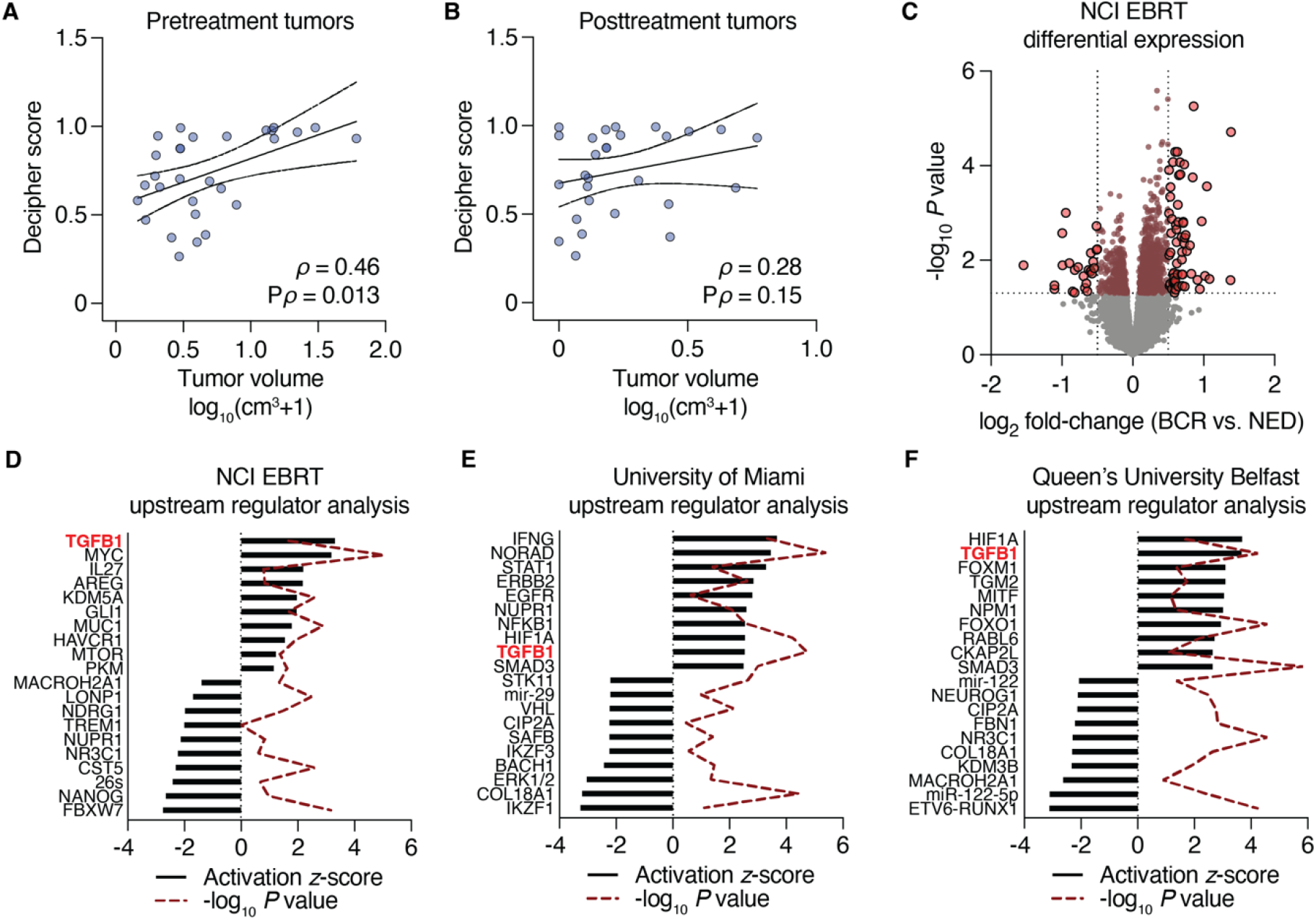
Transcriptomic analysis of baseline tumor biopsies. Patient-level MRI tumor volumes prior to treatment (**A**) or six months after RT (**B**) correlated with Decipher scores from a pre-selected biopsy (per patient). Nonparametric Spearman correlation analyses ρ values are shown with their respective P values. Volumes shown represent an added pseudocount that was log_10_-transformed. (**C**) Volcano plot depicting differentially expressed genes as determined using a linear mixed-effect model. Horizontal boundary depicts the P = 0.05 (unadjusted) cutoff, and vertical boundaries demarcate genes with a fold-change of at least ±2. For (**D-E**), differentially expressed genes (P < 0.05 unadjusted with no fold-change cutoff) comparing BCR vs. NED patients were analyzed with Ingenuity Pathway Analysis using the Upstream Regulator module. For (**F**), differentially expression genes (P < 0.01 unadjusted with no fold-change cutoff) were used. The 10 top and 10 bottom upstream regulators from each analysis are shown.

We extended our analyses to all 60 biopsy cores acquired and modeled differential expression as a function of outcome (BCR vs. NED) using a linear mixed-effect model to account for repeated and variable sampling across patients, modeling each patient as a random effect. This analysis yielded 1,083 differentially-expressed genes (P < 0.05, unadjusted), of which 83 genes were differentially expressed by at least 2-fold (**Fig. 3C**). Using all 1,083 genes in Ingenuity Pathway Analysis to determine upregulated pathways and master regulators enriched in BCR vs. NED cases, we identified TGF-β signaling as the top-most activated pathway (out of 58 activated upstream regulators) by upstream analysis (**Fig. 3D**).

To validate these findings, we similarly derived differential gene expression from two additional cohorts. The first cohort, from the University of Miami BLaStM trial, consisted of 21 patients with up to six baseline mpMRI/ultrasound-fusion targeted biopsies per patient sampled prior to the start of RT with RNA profiled using the Affymetrix Human Exon array. Three of these patients had experienced BCR at the time of analysis. Repeating the differential expression analysis using a linear mixed-effect model, we identified 1,993 genes that were differentially expressed (P < 0.05) between BCR and NED patients.

When applied to Ingenuity upstream regulator analysis, TGF-β emerged as the 9^th^-most (out of 130) activated regulator (**Fig. 3E**). The second validation cohort, from the Queen’s University Belfast, was comprised of 248 patients (56 with BCR) who had undergone conventional (templated) biopsy prior to RT with RNA profiled on the Affymetrix Metastatic Assay array [4]. We used a generalized linear model to determine differential gene expression between BCR and NED patients, as only a single biopsy represented each patient. We identified 1,557 differentially expressed genes, and Ingenuity again nominated TGF-β as significantly enriched, scoring as the second-most (out of 133) activated upstream regulator (**Fig. 3F**). Collectively, these results suggest that tumor cell biology underlying TGF-β signaling in an untreated prostate cancer setting is significantly associated with greater propensity to progress, manifesting clinically as a biochemical recurrence after RT.

### Genomic correlates of RT failure

We sought to determine whether somatic genomic alterations present at baseline were significantly associated with outcome following RT. We performed whole-exome sequencing (see **Fig. 2**) on 57 biopsy tumor samples (1-4 per patient, mean target coverage 118.2 ×) paired to matched benign DNA from tissue and/or buffy coat (mean target coverage 66.9 ×). Limiting our analyses to those genes nominated as significantly mutated in prostate cancer from previous cohort studies [5-7], no recurrent mutation of any single gene demonstrated enrichment in BCR vs. NED patients at P < 0.05. Notably, however, loss-of-function alterations to *SPOP* were exclusively observed in NED cases, consistent with this genotype’s association with favorable outcomes [8].

We thus restricted our analyses to biallelic inactivation (mutation and/or 1-2 copy number loss) events of tumor suppressors. Of the five genes that met this criteria, only two genes (*TP53* and *PTEN*) were altered in BCR cases (**Fig. 4A**). Biallelic alterations to *TP53* were enriched in BCR cases (P_adj_ = 0.046). Examining either a single selected core per patient (**Fig. 4B**) or all cores (**Fig. 4C**), we found that the number of *PTEN* or *TP53* alleles altered per tumor were positively associated with the probability of BCR. When a biopsy harbored at least three hits to *PTEN* and *TP53* alleles (such as hemizygous loss of *PTEN*, a mutation to *TP53* and deletion of the remaining *TP53* copy), this criteria was 100% specific for BCR cases when applied to pre-selected biopsy cores.

**Figure 4.**
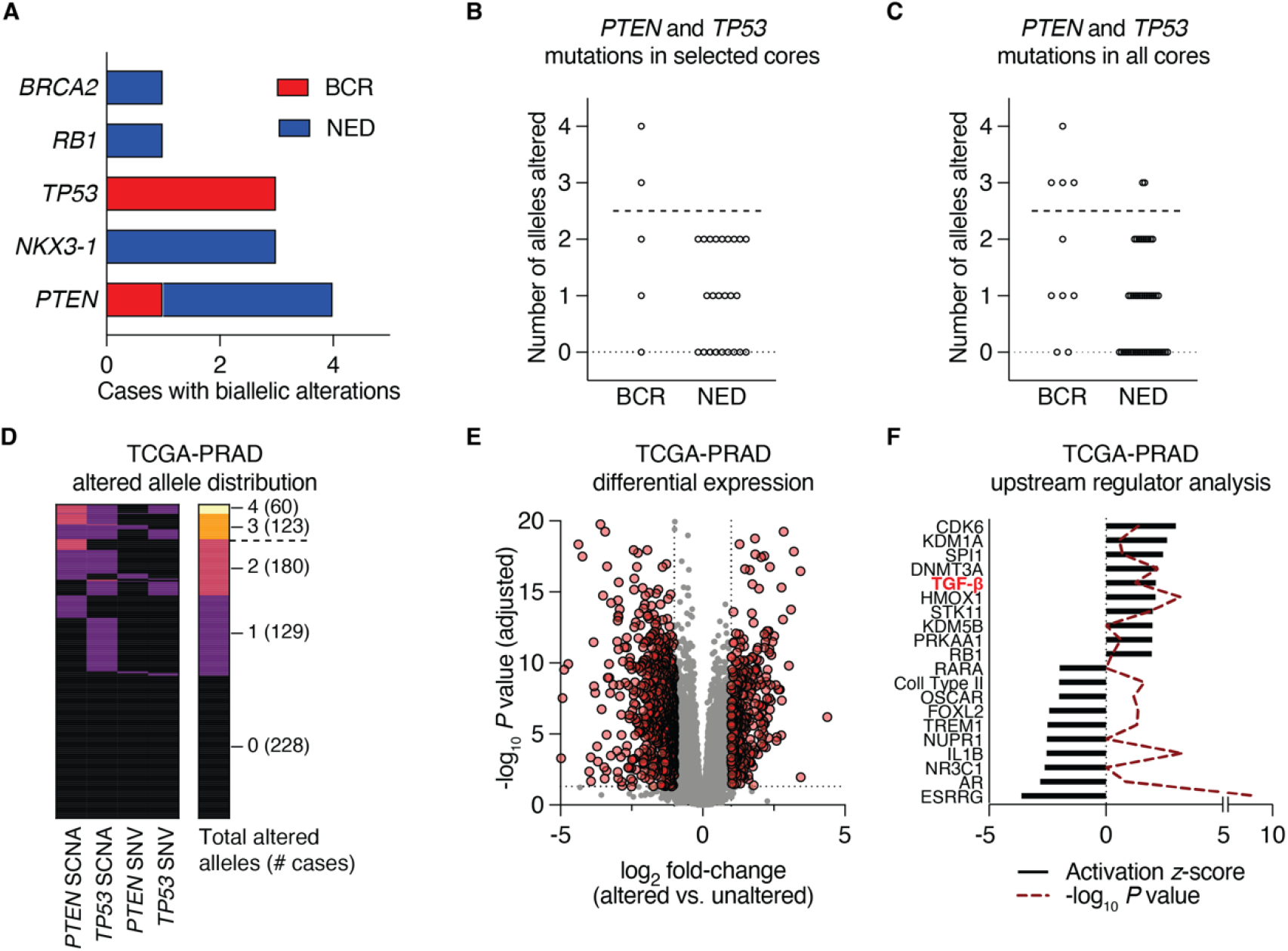
Biallelic tumor suppressor loss is associated with RT failure and increases in TGF-β signaling. (**A**) Frequency distribution of BCR and NED cases harboring biallelic losses to the indicated tumor suppressor genes. (**B-C**) Specifying a cutoff of three or four alleles altered (PTEN^-/-^; TP53^+/-^ or PTEN^+/-^; TP53^-/-^ or PTEN^-/-^; TP53^-/-^), samples above the dotted line pass this threshold when considering only selected cores (**B**) or all cores (**C**). (**D**) Distribution of cases from the prostate cancer TCGA (TCGA-PRAD) cohort with 0, 1, 2, 3, or 4 alleles of *PTEN* and *TP53* altered. The dotted line indicates the 3-allele cutpoint. (**E**) Volcano plot depicting differentially expressed genes as determined using a generalized linear model to compare TCGA-PRAD cases with 3-4 altered *PTEN* and *TP53* alleles vs. cases with 0-2 altered alleles. Horizontal boundary depicts the P = 0.05 (adjusted) cutoff, and vertical boundaries demarcate genes with a fold-change of at least ±2. (**F**) Genes with an average expression of 5 counts in 50% of the samples, a fold-change of at least ±2, with P_adj_ < 0.05 were analyzed with Ingenuity Pathway Analysis using the Upstream Regulator module. The 10 top and 10 bottom upstream regulators are shown.

Although not treated by radiation therapy, the prostate cancer TCGA [5] represents a large number of clinically similar untreated localized prostate tumors. Therefore, we employed the *PTEN* and *TP53* allele cut-off to this larger cohort of cases, stratifying the prostate cancer TCGA cohort based on manually curated calls for *PTEN* and *TP53* using the same computational pipeline as our own analysis. This resulted in 55 tumors with 3 or 4 *PTEN* and *TP53* alleles altered and 443 tumors with 0, 1 or 2 alleles altered (**Fig. 4D**). To test whether this genomic classification recapitulated a similar phenotype as we observed in tumors prior to RT, we performed differential gene expression on matched RNA-seq data available for those cases. After comparing the more altered tumors (3-4 alleles) to the less altered tumors (0-2 alleles), we selected 1,067 transcripts differentially expressed by at least 2-fold with an adjusted P value cutoff of 0.05 (**Fig. 4E**). Similar to our prior analyses, Ingenuity upstream regulator analysis nominated TGF-β signaling as a pathway with the 5^th^-highest activation *z*-score (**Fig. 4F**).

Taken together, these findings suggest that severe disruptions to *PTEN* and *TP53* function in untreated prostate tumors may be independently associated with elevated TGF-β activity and may serve as a genomic biomarker of tumors with propensity to resist RT and/or ADT.

### Independent phenotypic convergence to TGF-β activity

As we have observed an association of tumor transcriptomics with TGF-β signaling in 3 cohorts of RT cases as well as prostate TCGA when selecting cases matching the genomic profile of tumors that fail RT, we next asked whether MRI volumes, which independently were associated with RT failure, also tracked with TGF-β. We applied a linear mixed-effect model and modeled each patient as a random effect to identify genes that correlated with greater pretreatment or posttreatment volumes. Although many canonical tumorigenic pathways were amongst the top upstream regulators nominated by Ingenuity Pathway Analysis, TGF-β was the 17^th^-highest (pretreatment, **Fig. 5A**) and 12^th^-highest (posttreatment, **Fig. 5B**), out of 128 and 70 activated pathways, respectively. As this association was independent of RT outcome (although volumes were prognostic of RT failure, see **Fig. 1**), it suggests that TGF-β activity may be an independent predictive variable for tumor volume in this setting.

**Figure 5.**
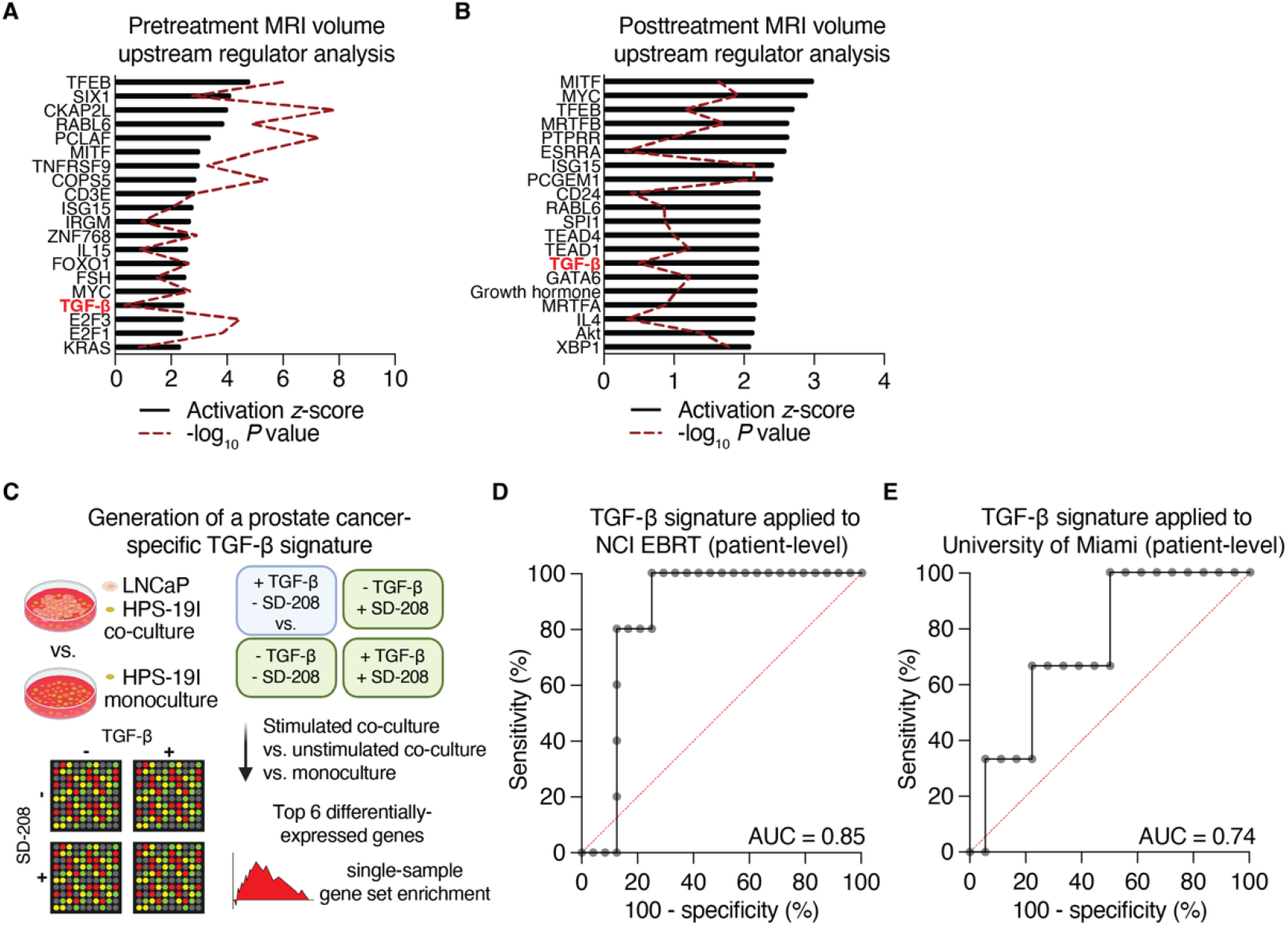
TGF-β activity is independently associated with outcome following RT. (**A-B**) Gene expression on biopsies (*n*=56 which excludes biopsies targeted to seminal vesicles) sampled from 38 MRI lesions were correlated with those lesions’ pretreatment (**A**) or posttreatment (**B**) volumes using a linear mixed-effect ranked-outcome model. All genes with unadjusted correlation P values less than 0.05 were analyzed with Ingenuity Pathway Analysis using the Upstream Regulator module. The top 20 activated upstream regulators are shown. (**C**) Schematic for building a prostate cancer-specific TGF-β activation signature using HPS-19I stromal cells with and without co-culture of LNCaP epithelial cells, ± TGF-β stimulation, ± TGF-β receptor inhibition (SD-208). (**D-E**) Receiver operating characteristic curves for the prostate cancer-specific TGF-β signature to distinguish BCR from NED cases in the NCI EBRT (**D**) and University of Miami (**E**) cohorts when applied to the pre-selected biopsy core from each case.

These findings of TGF-β activity are based on Ingenuity’s curated pathway database, in which input is comprised of differentially-expressed gene list from a cohort of patients. To develop a signature more amenable to determining individualized risk, we started by generating a prostate cancer-specific TGF-β signature based on microarray data from LNCaP and HPS-19I cells [9]. This gene signature is comprised of the top six differentially expressed genes between LNCaP prostate cancer epithelial cells grown in co-culture with HPS-19I cells that were stimulated TGF-β, versus 5 control conditions: LNCaP/HPS-19I co-culture without TGF-β stimulation, LNCaP/HPS-19I with SD-208 TGF-β receptor inhibitor (with or without TGF-β simulation), and HPS-19I cells grown alone (with or without TGF-β stimulation) (**Fig. 5C**).

As described in Methods, the top six differentially-expressed genes identified by this approach were capable of discriminating between BCR and NED cases in the NCI EBRT cohort with an AUC of 0.85 (**Fig. 5D**). The Queen’s University Belfast cohort profiled on the Affymetrix Metastatic Assay array did not contain all of genes in our final signature, and therefore we could not test its utility in that group.

However, when applied our prostate cancer-specific TGF-β signature to the University of Miami cohort, test performance AUC equal to 0.77 (**Fig. 5E**). These findings indicate that TGF-β is independently associated with the development of localized intermediate and high risk prostate cancer and its elevated activity is prognostic for failure following EBRT plus ADT.

## DISCUSSION

In the current study, we conducted a radiogenomic analysis of tumors from patients with intermediate, high, and very high risk localized prostate cancer treated with radiotherapy and ADT, integrating clinical, imaging, and pathologic data with genomic profiles. We identified TGF-β as a key upstream regular of gene expression in patients destined to develop BCR after EBRT ± ADT in several distinct cohorts. Separately, predicted TGF-β activation was associated with biallelic losses to *PTEN* and *TP53* and correlated with tumor volumes before and after EBRT ± ADT. We developed a novel prostate cancer-specific TGF-β gene signature that was prognostic for biochemical recurrence after EBRT ± ADT. Our work thus identifies activation of TGF-β signaling as key negative prognostic indicator in untreated, localized prostate cancer.

In localized prostate cancer, TGF-β has canonically been associated with tumor-suppressive and antiproliferative phenotypes [10]. However, reports from our group and others identified TGF-β signaling through SMAD proteins as a putative mechanism of progression from lower to higher grade prostate cancer [11-13]. The dichotomy of TGF-β signaling playing a suppressive role in premalignant cells and a stimulatory role in cancer cells has been attributed to contextual determinants that promote TGF-β mediated epithelial mesenchymal transition (EMT) while simultaneously restricting the cytostatic effects of TGF-β [14, 15]. However, the importance of TGF-β signaling on treatment response, especially treatment combinations involving radiotherapy combined with ADT, is less certain. A TGF-β transcriptional program in pretreatment tumor biopsies has previously been identified as a marker of poor response to neoadjuvant-intense androgen deprivation therapy [16]. TGF-β signaling has been implicated in EMT of prostate cancer cells, which, in turn, has been implicated in treatment related neuroendocrine differentiation and therapeutic resistance to androgen deprivation therapy and radiotherapy [17]. Thus, whether the TGF-β transcriptional program in our series is a marker for resistance to the radiotherapy and/or ADT component of treatment is uncertain.

A compelling component of the present study is the linkage of a *TP53* and *PTEN* mutational profile, which has known negative prognostic importance, to an activated TGF-β transcriptional signature in independent localized prostate cancer datasets. Losses to *TP53* and *PTEN* are associated with EMT, the emergence of castration resistance, and resistance to ADT in mouse models of prostate cancer and numerous patient cohorts [18-21]. Our finding that disruption of 3 or 4 *TP53* and *PTEN* alleles in untreated prostate tumors serves as a causative factor in an activated TGF-β transcriptional signature is consistent with proposed models in which TGF-β can either directly activate AKT independent of SMADs to promote proliferation or alternatively cooperate with *TP53* loss to re-engage developmental pathways, including EMT, to facilitate metastasis [22-24]. As inhibition of TGF-β signaling has been shown to delay prostate cancer progression to castration resistance *in vitro* and *in vivo*, a tantalizing opportunity arising from the present study may be the incorporation of systemic therapies targeting TGF-β, including TGF-β traps, in the backbone of EBRT and ADT in the setting of high risk localized prostate cancer [25, 26].

Molecular classifiers such as the Decipher Genomic Classifier, the Oncotype DX Genomic Prostate Score, and the Prolaris Cell Cycle Progression test were designed to improve stratification beyond standard risk grouping schemas [27]. The Decipher test, which was originally designed for prediction of metastatic potential following radical prostatectomy, has demonstrated utility in other disease settings, including assessing clinical benefit from definitive therapy for localized disease and radiotherapy following prostatectomy [28-30]. Because the Decipher test uses a genome-wide microarray platform, the same technical processes have led to development of additional signatures and scores relevant to prostate cancer treatment, such as an ADT resistance score and the PORTOS score for benefit from post-prostatectomy radiotherapy [31, 32]. Common to most of these signatures was the use of machine learning for unbiased feature selection to optimize assay performance, but this has yielded little insight into other targetable processes and pathways that contribute to local or metastatic recurrence after treatment. By contrast, we generated a novel TGF-β signature using *in vitro* models of TGF-β activity, in which selected features are consistent with known patterns of TGF-β receptor and SMAD protein activation. Although we utilized multiply-sampled tumors to quantify differentially expressed genes which led us to identify TGF-β as a prognostic biomarker of BCR, our novel TGF-β signature independently distinguished BCR from NED with 80% accuracy. Our index cohort was profiled using the same microarray platform as Decipher tests and we designed our novel TGF-β signature to operate on data processed using that platform; this will enable the TGF-β signature to be validated and applied in wider settings as part of future planned investigations.

This study has several limitations. Prostate MRI is often used for the purposes of staging prior to treatment but is rarely used in response assessment in the modern care of patients with localized prostate cancer treated with EBRT. Consequently, although we completed a comprehensive radiogenomic analyses on a subset of patients in our index cohort, a suitable confirmatory cohort with similar radiologic data could not be identified. Second, although we used WES and gene expression data from clinically similar patients with untreated localized prostate cancer from the TCGA to validate the link between biallelic losses to *PTEN* and *TP53* with activation of TGF-β signaling, we could not identify a appropriate validation cohort of tumor biopsies with paired WES and gene expression data from patients treated with EBRT ± ADT. Finally, the TGF-β gene signature we developed in this study predicts for biochemical recurrence and metastatic failure in the index cohort, but the validation cohorts we tested only catalogued the presence of BCR. Thus, whether this signature not only predicts for BCR but also the pattern of failure requires further validation in suitable cohorts.

By performing comprehensive genomic and transcriptomic analysis of tumors using modern technologies, including a clinically validated array platform in commercial use, in a cohort of patients with extended follow-up we have linked the activation of TGF-β signaling to biallelic loss of *PTEN* and *TP53*, and eventual BCR after EBRT ± ADT. This study also links tumor response as measured by MRI to the same signaling pathways that correlate to biochemical recurrence, suggesting that these processes are collectively linked to a dominant pattern of TGF-β signaling. Collectively, our findings provide biological insight in the biology underlying predisposition to BCR following EBRT ± ADT and open additional avenues for investigation to target TGF-β signaling in the high risk localized prostate cancer setting.

## PATIENTS AND METHODS

### Study Approval

This trial was approved by the Institutional Review Board of the Center for Cancer Research, NCI (Bethesda, MD; ClinicalTrials.gov Identifier: NCT01834001). This study has been conducted in accordance with ethical principles that have their origin in the Declaration of Helsinki and are consistent with the International Council on Harmonization guidelines on Good Clinical Practice, all applicable laws and regulator elements, and all conditions required by a regulatory authority and/or institutional review board. Written informed consent was obtained from all patients prior to performing study-related procedures in accordance with federal and institutional guidelines.

### Study Design

In this study, men with newly diagnosed intermediate or high risk localized prostate cancer underwent targeted research biopsy and multiparametric MRI (mpMRI) prior to standard of care definitive external beam radiotherapy (EBRT) with or without androgen deprivation therapy. Eligible patients had histologically confirmed intermediate or high risk prostate cancer with an MRI visible lesion and no prior systemic therapy or local therapy for prostate cancer. Patients with contraindication to prostate biopsy or contrast enhanced mpMRI were excluded. Whole blood was acquired prior to the initiation of therapy.

Androgen deprivation therapy was typically delivered for 2 months prior to initiation of EBRT. The total duration of androgen deprivation therapy depended on risk grouping and tolerance. Patients underwent a second mpMRI at 6 months after completion of radiation treatment. Patients were followed clinically with a serum PSA and physical examination every 3 months for the first two years, every 6 months from year 2 through 5, and then yearly. Biochemical recurrence was defined as a rise in PSA of 2 ng/mL or more above the post treatment nadir [33].

### mpMRI

All patients underwent 3T mpMRI prior to any treatment and 6 months after completion of radiation.

Multiparametric MRI included T2-weighted (T2W), diffusion-weighted (DWI), and dynamic contrast-enhanced (DCE) imaging with the use of an endorectal coil or phased array surface coil. MRI images were reviewed prospectively by one prostate dedicated radiologist with more than 10 years of individual experience using standard institutional image interpretation method at both baseline and 6 months post-treatment follow up MRIs (PIRADS was not used since the study accrual started before release of PIRADS and interpretation also included post-treatment MRIs). For both baseline and follow-up MRIs, lesions were manually contoured on T2W using mpMRI pulse sequence with commercially available software (version 6.6.10; MIM), which enable spatial referencing to images from all other sequences to ensure lesion features (e.g., lesion volumes and apparent diffusion coefficient (ADC) values) were encompassed in entirety within the volume of interest.

### Prostate tissue processing

Diagnostic and research targeted biopsies from MRI-visible prostate lesions acquired prior to any therapy underwent standard pathologic evaluation for presence and grading of tumor by a genitourinary specialized anatomic pathologist (H.S.). Serial sections (5 μm) of formalin-fixed, paraffin-embedded biopsy tissues were stained with hematoxylin and eosin, stained with antibodies via immunohistochemistry (IHC), or cut as ribbons for nucleic acid extraction.

For IHC, slides were stained using validated protocols with antibodies ERG and PIN-4 using manual staining approaches as previously described [18]. Glass slides were baked for 15 min at 60 °C, except for PIN-4 staining which was baked overnight at 45 °C. Following deparaffinization in xylenes and rehydration through graded alcohols, antigen retrieval was performed using a NxGen Decloaker (Biocare Medical) at 110°C for 15 minutes in Diva Decloaker (BioCare Medical; DV2004MX). Sections were blocked with hydrogen peroxide (Sigma Aldrich, Cat# 216763) for 5 min, blocked with Background Sniper (Biocare Medical, Cat# BS966) for 10 min for PIN-4 or VectaStain Elite ABC HRP kit (Vector Laboratories, Cat# PK-6101) for ERG, and incubated with primary antibody for overnight at 4 °C (ERG) or 1 h (PIN-4). 50 μl of antibody solutions were prepared for anti-ERG (Abcam, Cat# ab92513, 1:500 into SignalStain diluent) and PIN-4 cocktail (Biocare Medical, Cat# PPM225DS, ready-to-use). SignalStain diluent was from Cell Signaling. Secondary labeling was performed using Mach 2 Double Stain for PIN-4 (Biocare Medical, Cat# MRCT525) or the VectaStain Elite ABC HRP kit for ERG for 30 min. Avidin–biotin complexing was then performed for 30 min for ERG. Colorimetric detection was achieved using DAB Peroxidase HRP (Vector Laboratories, Cat# SK4100) for ERG or Vulcan Red Fast Chromogen (Biocare Medical, Cat# FR805) with Betazoid DAB (Biocare Medical, Cat# BDB2004) for PIN-4. Counterstaining was performed using Mayer’s Hematoxylin Solution (Sigma Aldrich, Cat# MHS16). PIN-4 stained slides were air-dried. ERG-stained slides were dehydrated through graded alcohol and cleared in xylenes. Slides were mounted using Permount (Thermo Fisher).

Prior to performing molecular analyses, one core was pre-selected from each patient as representative of that tumor, using criteria that would be applied if sending a single sample per patient for commercial molecular diagnostics. Amongst all biopsy cores with tumor, the sample with the highest Gleason Grade Group was chosen. In the case of tie between two or more biopsies, the core with the greatest percentage of tumor belonging to the highest grade group was selected.

### Gene expression analyses

Total RNA was extracted from 1-3 FFPE ribbons of biopsy tissues containing tumor, using the RNeasy FFPE Kit (Qiagen, Cat# 73504) with minor modifications to the manufacturer’s protocol. First, after incubation with deparaffinization solution (Qiagen, Cat# 19093), the sample was allowed to cool for 3 minutes to room temperature. Second, the volume of Proteinase K was increased to 40 μl. Finally, the initial lysis incubation at 56 °C was extended to two hours with shaking at 1000 rpm. After determining yield using Qubit RNA Assay kits (Invitrogen, Cat# Q32852), 25-100 ng of RNA were amplified into fragmented and biotinylated cDNA using the Ovation FFPE WTA kit (NuGEN, Cat# 3403-60) and the Encore Biotin Module (NuGEN, Cat# 4200-60) as previously described [29]. cDNA was hybridized to Human Exon 1.0 ST microarrays (Affymetrix, Cat# 900651) using an Affymetrix GeneChip Fluidics Station and scanned using an Affymetrix GeneChip Scanner at the NCI Genomics Technology Laboratory to generate CEL files. CEL files for the University of Miami cohort were provided by Dr. Alan Pollock. CEL files for the Queen’s University Belfast cohort [4] were downloaded from GEO (Accession ID GSE116918).

To identify differentially-expressed genes, CEL files were first normalized using RMA using Bioconductor. For Affymetrix Human Exon arrays, gene symbols were summarized from probe values using the pd.huex10st.hs.entrezg platform definition available from BrainArray. For Affymetrix Metastatic Assay arrays, a platform definition was built using the ADXPCv1a520642 probe definitions and probe sequences available through GEO (https://www.ncbi.nlm.nih.gov/geo) and Almac Diagnostics with pdInfoBuilder for Bioconductor. Limma [34] was used with the lmfit linear model for comparing a single biopsy per case. For considering multiple biopsies per case, the voom model was first used to identify mean variance between cores from the same patient before applying weights into an empirical Bayes moderation. With voom, patient identifiers were used as blocking factors to achieve a linear mixed-effect model.

Differentially-expressed genes from prostate TCGA cases harboring *PTEN* and *TP53* mutations (see DNA somatic alteration analyses, below) were determined first by downloading the paired-end RNA-seq FASTQ files of tumor cases from the NCI Genomic Data Commons (https://gdc.cancer.gov) under dbGaP Authorized Access Research Project ID 9940. FASTQ pairs were aligned to hg19 and summarized as gene-level counts using RSEM [35] version 1.3.2 as a wrapper around STAR [36] version 2.7.0f. Differentially-expressed genes were identified using a generalized linear model with DESeq2 for R [37].

Candidate upstream regulators were identified using Ingenuity Pathway Analysis. Gene lists delivered from differential expression analyses were uploaded with their corresponding log_2_ fold-change and P values. The upstream regulator module of the core pathway analysis reports individual molecules, pathways and master regulators with an activation *z* score and P value of overlap of the submitted gene list with Ingenuity’s curated gene networks.

Gene level summaries from Agilent 4×44K microarrays of LNCaP and HPS-19I cultures [9] were downloaded from GEO (Accession ID GSE51624). Differential expression was computed using limma, separately for the co-cultures and the stroma alone. The model for co-culture differential gene expression consisted of the TGF-β stimulated LNCaP/HPS-19I co-culture without inhibitor against five equally-weighted negative conditions (DMSO only, TGF-β plus SD-208, and SD-208 only from co-culture, and TGF-β stimulated or vehicle treated HPS-19I cells alone) passed to lmfit. All genes with adjusted P value less than 0.1 were ranked by log2(fold-change). This process selected for genes enriched only in epithelial cells to and generate a prostate cancer-specific gene list of TGF-β activity.

TGF-β signature scores were calculated by processing the prostate cancer-specific gene list and normalized expression value with GSVA [38] for R. GSVA was performed using method=“ssgsea” with tau set to 0.25. The single-sample gene set enrichment score for TGF-β activity was used directly with the roc and auc functions in R to perform receiver operating characteristic analysis and determine the area under the ROC curve, respectively. The final list of genes for use in signature was refined in the NCI EBRT discovery cohort by iteratively running the top *N* genes (starting at three genes) and selecting the local maximum AUC value for this cohort. At this point, the gene set was locked and used for validation in the University of Miami cohort.

### DNA somatic alteration analyses

Tumor was enriched from FFPE tumor baseline biopsy sections by microdissection with a scalpel using hematoxylin-stained slides under a brightfield microscope, using reference slides stained with H&E and PIN-4 to identify tumor regions marked by a pathologist (H.S.). Additional regions of benign tissue were acquired for 28 of the 29 patients. Up to 9 serial slides (cut at 5 μm depth) were pooled per sample. DNA was isolated from these tissue scrapes using the QIAamp DNA FFPE Tissue Kit (Qiagen, Cat# 56404) with minor modifications to the manufacturer’s protocol. First, the initial lysis step at 56 °C was extended to overnight, and the final elution step was performed twice to increase yield. DNA was extracted from buffy coat collected in K_2_EDTA tubes prior to treatment using the QIAamp DNA Blood Mini Kit (Qiagen, Cat#51104) as per the manufacturer’s protocol. Recovery of double-stranded DNA was confirmed using the Qubit dsDNA Assay Kit (Invitrogen, Cat# Q32851) and the Quant-iT PicoGreen dsDNA Assay Kit (Invitrogen, Cat# P11495).

Whole-exome sequencing libraries were prepared from 50-1000 ng of DNA isolated from buffy coat and tissue samples using the SureSelect XT Low Input Reagent Kit with the Human All Exon v6 Target Enrichment Baits (Agilent, Cat# G9707K). Paired-end libraries were sequenced on 31 lanes of a HiSeq 4000 (Illumina) with 2 × 100 cycles and processed into FASTQ files using bcl2fastq version 2.17 (Illumina). Reads were trimmed to remove adaptor sequences using SurecallTrimmer version 4.0.1 (Agilent) and aligned by read group to GRCh38 (including decoy contigs) using BWA version 0.7.17. Lane-level alignments were duplicate-marked and quality score recalibrated using GATK version 4.2.6.1, merged per-sample, and duplicate-marked a second time.

Germline mutations to coding genes were identified using GATK HaplotypeCaller, first generating a per-sample GVCF then then genotyping it using GATK GenotypeGVCFs. Indels and SNPs were separately calibrated using GATK VariantRecalibrator and GATK ApplyVQSR using the Mills & Gold 1000 Genomes, HapMap version 3.3, dbSNP version 138 and 1000 Genomes Phase 1 SNPs as ground truth references. Mutations were annotated using ANNOVAR [39] build 2020-06-08.

A unified set of point mutations and indels were established by intersecting the final calls of MuTect2 and Strelka. To run MuTect2, GATK MuTect2 version 4.2.6.1 was first run in tumor-only mode against each benign FFPE tissue BAM for the intervals of bait targets. A panel-of-normals was created from the resultant VCF files using GenomicsDBImport and CreateSomaticPanelOfNormals. Then, MuTect2 was run a second time in paired mode using each tumor BAM paired to its buffy coat-derived normal. Raw calls were filtered against tumor-in-normal contamination (using GetPileupSummaries and CalculateContamination) and read orientation artifacts (using LearnReadOrientationModel and FilterMutectCalls). To run Strelka (IIlumina), BAM files were first local indel (left) realigned using GATK LeftAlignIndels. Manta was run in paired mode using default parameters of the Manta workflow for exome sequencing. The indelCandidates detected by Manta were passed to Strelka and processed using the Strelka workflow’s default parameters for exome sequencing. Somatic indels and SNVs were annotated using VAtools. Tumor-mutation annotated VCF files from MuTect2 and Strelka were intersected, and common somatic mutations for each tumor sample were further annotated using ANNOVAR build 2020-06-08.

Allele-specific somatic copy number alterations were identified at the gene and chromosome level by manually comparing the output of GATK version 4.2.6.1, CNVkit version 0.9.9 [40] and facetsSuite version 2.0.0 [41] and deriving a curated and unified set of somatic copy number gains and losses for each tumor sample. To run GATK, bait intervals were pre-processed and annotated for mappability and GC content using PreprocessIntervals and AnnotateIntervals. Read counts from each tumor or FFPE benign BAM file were obtained using CollectReadCounts on regions covered by baits. A panel-of-normals using just thee FFPE benign read counts were created using CreateReadCountPanelOfNormals, and the panel-of-normals was used to denoise tumor-level read count files to create denoised copy ratios using DenoiseReadCounts. B-allele frequencies were collected against all captured intervals using CollectAllelicCounts on all tumor BAMs and buffy coat derived normal BAMs. Tumor denoised copy ratios, tumor allelic counts and benign allelic counts were jointly modeled with ModelSegments to derive intervals of somatic copy number gain or loss using default parameters to produce a somatic copy number SEG file.

To run CNVkit, accessible regions of GRCh38 at 5000-base resolution were defined using cnvkit access, and the accessible regions were then binned based on the BED file of the exon capture bait file using cnvkit autobin. The bin size for on-and off-target (target or antitarget) regions were annotated to target and antitarget BED files against refFlat (downloaded from UCSC goldenPath). CNVkit was then run on the benign FFPE tissue BAM files first to assess coverage for targeted and nontargeted regions filtering for reads with a minimum mapQ of 10 using cnvkit coverage. The target and antitarget coverage CNN files were then combined to make a project-specific reference, also locking the sample sex to male. Next, tumor BAM files were similarly processed using cnvkit coverage for targeted and non-targeted regions, which were then passed to cnvkit fix to determine log-ratio coverage against the reference, and then segmented using cnvkit segment into CNS files. Allele-specific events were determined by passing tumor alterations in VCF files from MuTect2 to cnvkit call, which modified each segmented sample to account for allele-specific alterations with additional arguments --drop-low-coverage and --filter cn. The final B-allele frequency annotated CNS files were converted to a SEG file using cnvkit export. facetsSuite was run using the provided Rscript wrapper, which took SNP pileup files (generated using the provided snp-pileup Rscript wrapper and left-aligned tumor BAMs paired with benign buffy coat BAMs) and generated SEG files along with purity and ploidy estimates.

Broad and focal areas of chromosome loss and gains were determined using GISTIC2. SEG files generated by GATK, CNVkit and facetsSuite were separately passed to GISTIC2 using the following arguments: -b gistic -genegistic 1 -broad 1 -brlen 0.5 -conf 0.90 -armpeel 0 -savegene 1 -gcm extreme - maxspace 10000 -maxseg 10000 -remove_x 0 -cap Inf. Arm-level gains and losses inferred by GISTIC2 were manually inspected across all three copy number SEG inputs and only consensus calls were reported.

498 tumor-normal pairs of whole-exome sequencing BAM files from the prostate cancer TCGA cohort were downloaded from the NCI Genomic Data Commons. FASTQ files were recovered from BAM files using PICARD samtofastq with OUTPUT_PER_RG set to TRUE. Lane-level FASTQ pairs were then realigned to the same version of GRCh38 as our prior alignments with the same duplicate-marking and quality score recalibration pipeline as described above. Lane-level BAM files were merged, and duplicate marked a second time. Detection of somatic alterations was performed exactly as described above. For determining oncogenic function of alterations identified in the NCI and TCGA cohorts, somatic events to cancer-related genes were queried against OncoKB [42]. Only alterations deemed “likely oncogenic” and “oncogenic” were reported.

### Statistical analyses

Statistical analyses were performed with GraphPad Prism version 8 (GraphPad Software Inc., San Diego, CA, USA), Microsoft Excel for Mac version 16 (Microsoft Corporation, Redmond, WA, USA), and R version 2.4 (R Core Team 2023). Comparisons of single factors between biochemical recurrence (BCR) patients and no evidence of disease (NED) patients were performed using Mann-Whitney *U* tests. Null hypothesis tests of associations (enrichments) between response and individual dichotomous factors were performed using a two-sided Fisher’s exact test at the per-patient level. Associations between factors were measured using Spearman correlations. The significance of differential gene expression measures was determined using generalized linear models and linear-mixed effect models.

The accuracy of gene set enrichment tests to predict dichotomized outcomes was examined using receiver operating characteristic curves.

## Data Availability

The data underlying this article have been deposited in Database of Genotypes and Phenotypes (dbGaP) and Gene Expression Omnibus (GEO) at https://www.ncbi.nlm.nih.gov/gap/ and https://www.ncbi.nlm.nih.gov/geo/, respectively, and can be accessed with phs001821.v1.p1 (dbGaP) and GSE129274 (GEO).

https://www.ncbi.nlm.nih.gov/geo/

## DECLARATIONS PAGE

### Ethics approval and consent to participate

The collection and analysis of tissue from patients with localized prostate cancer was approved by the Institutional Review Boards of the National Institutes of Health (NCT01834001, “Imaging Studies to Check the Local Response of Prostate Cancer to Radiation Therapy”, PI: Deborah Citrin) and the University of Miami Sylvester Cancer Center (NCT02307058, “Randomized MRI-Guided Prostate Boosts Via Initial Lattice Stereotactic vs Daily Moderately Hypofractionated Radiotherapy (BLaStM)”, PI: Alan Pollack). All patients provided informed consent before participating. This research was conducted in accordance with the principles of the Declaration of Helsinki.

### Consent for publication

Not applicable.

### Authors’ contributions

<u>Data acquisition:</u> S.Y.T., H.Z., S.A.H., N.V.C., R.S., B.T.

<u>Methodology:</u> A.T.K., U.S., S.A.H., Y.L., B.J.W., P.P., R.S., E.D., A.P., B.T., A.G.S., D.E.C.

<u>Reagents:</u> P.P., B.J.W, R.S., A.G.S., D.E.C.

<u>Analysis:</u> A.T.K., U.S., S.Y.T., H.A.S., S.A.H., Y.L., R.S., E.D., B.T., A.G.S., D.E.C.

<u>Manuscript:</u> All authors

<u>Supervision:</u> A.G.S., D.E.C.

<u>Accessed and verified all underlying data:</u> A.G.S., D.E.C.

## Acknowledgments

The authors gratefully acknowledge the patients and the families of patients who contributed to this study. The authors thank Prof. Suneil Jain (Queen’s University Belfast) for his assistance in acquiring platform definition files to re-process the Affymetrix Metastatic Assay array data. Portions of this work utilized the computational resources of the NIH HPC Biowulf cluster.

## LIST OF ABBREVIATIONS

ADT: Androgen deprivation therapy
AR: Androgen receptor (gene, protein)
BCR: Biochemical recurrence
DCE: Dynamic contrast enhancement
DWI: Diffusion-weighted imaging
EBRT: External beam radiation therapy
mpMRI: mutiparametric magnetic resonance imaging
NCI: National Cancer Institute
NED: No evidence of disease
PSA: Prostate specific antigen
PTEN: Phosphatase and tensin homolog (gene, protein)
RT: Radiotherapy
T2W: T2-weighted
TGFβ: Transforming growth factor beta (gene, protein)
TP53: Tumor protein 53 (gene, p53 protein)

