## Supplementary Information for "Radiogenomic profiling of prostate tumors prior to external beam radiotherapy converges on a transcriptomic signature of TGF-β activity driving tumor recurrence"

Anson T. Ku *et al.*

- Supplementary Figures

**SUPPLEMENTARY FIGURES**

**Supplementary Figure 1**

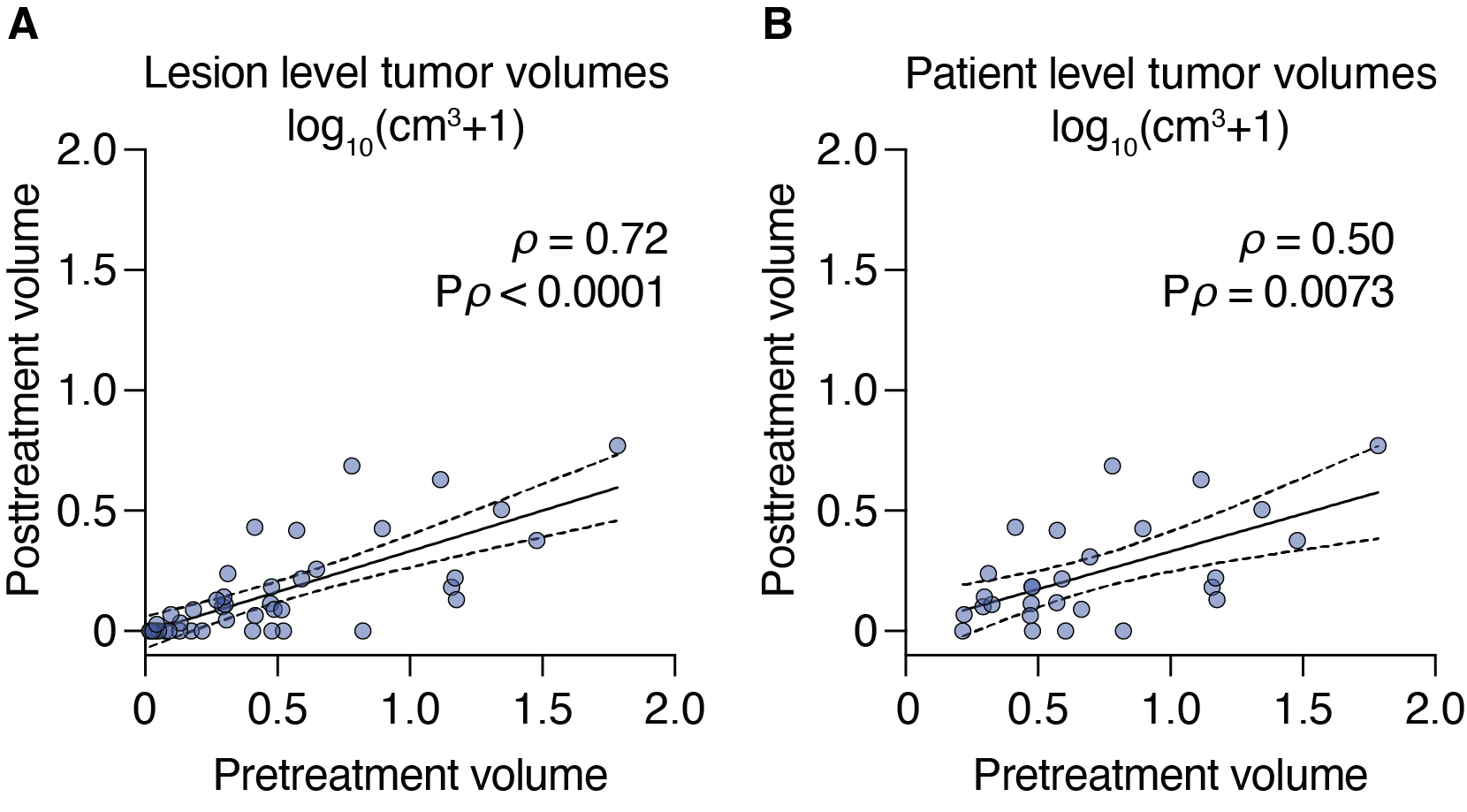

**Supplementary Figure 1. Correlations between pre- and posttreatment MRI tumor volumes.** Data is shown for individual lesions (A) and all lesions added together for a per-patient value (B). Nonparametric Spearman correlation analyses *ρ* values are shown with their respective P values. Volumes shown represent an added pseudocount that was log_10_-transformed for better visualization of posttreatment lesion volumes with volumes of zero cm^3^. Lines and bands represent linear regression lines and 95% confidence intervals, respectively.

**Supplementary Figure 2**

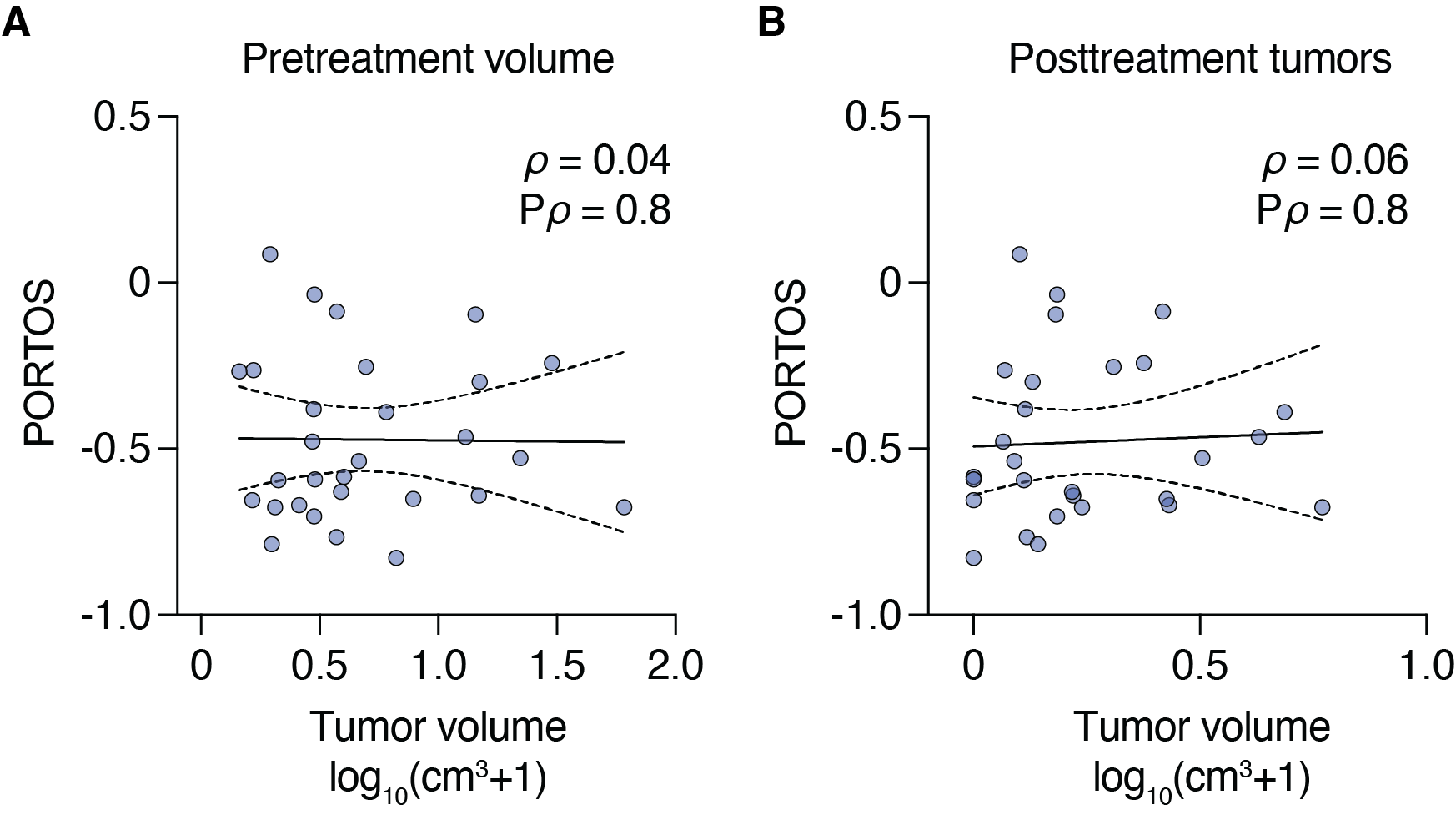

**Supplementary Figure 2. Correlations between MRI tumor volumes and PORTOS.** Data is shown for individual lesions (A) and all lesions added together for a per-patient value (B). Nonparametric Spearman correlation analyses *ρ* values are shown with their respective P values. Volumes shown represent an added pseudocount that was log_10_-transformed for better visualization of posttreatment lesion volumes with volumes of zero cm^3^. Lines and bands represent linear regression lines and 95% confidence intervals, respectively.
